# GlioMod: Spatiotemporal-Aware Glioblastoma Multiforme Tumor Growth Modeling with Deep Encoder-Decoder Networks

**DOI:** 10.1101/2022.11.06.22282010

**Authors:** Rishab K. Jain, Abhinav Gupta, Wael H. Ali, Pierre F. J. Lermusiaux

## Abstract

Glioblastoma multiforme is an aggressive brain tumor with the lowest survival rate of any human cancer due to its invasive growth dynamics. These dynamics result in recurrent tumor pockets hidden from medical imaging, which standard radio-treatment and surgical margins fail to cover. Mathematical modeling of tumor growth via partial differential equations (PDE) is well-known; however, it remains unincorporated in clinical practice due to prolonged run-times, inter-patient anatomical variation, and initial conditions that ignore a patient’s current tumor. This study proposes a glioblastoma multiforme tumor evolution model, GlioMod, that aims to learn spatiotemporal features of tumor concentration and brain geometry for personalized therapeutic planning. A dataset of 6,000 synthetic tumors is generated from real patient anatomies using PDE-based modeling. Our model employs image-to-image regression using a novel encoder-decoder architecture to predict tumor concentration at future states. GlioMod is tested in its simulation of forward tumor growth and reconstruction of patient anatomy on 900 pairs of unseen brain geometries against their corresponding PDE-solved future tumor concentrations. We demonstrate that spatiotemporal context achieved via neural modeling yields tumor evolution predictions personalized to patients and still generalizable to unseen anatomies. Its performance is measured in three areas: (1) regression error rates, (2) quantitative and qualitative tissue agreement, and (3) run-time compared to state-of-the-art numerical solvers. The results demonstrate that GlioMod can predict tumor growth with high accuracy, being 2 orders of magnitude faster and therefore suitable for clinical use. GlioMod is provided as an open-source software package, which includes the synthetic tumor data generated from the patients in our study.

## 1 Introduction

Glioblastoma multiforme (GBM) is the most aggressive brain tumor and deadly human cancer, with a 5-year survival rate of just 5.1% [1, 2]. GBM is conventionally treated with surgical resection followed by chemoradiotherapy [3]. However, due to the invasive and diffusive nature of GBM, tumor cells are often left behind after treatment resulting in tumor recurrence [4]. These recurrence-inducing pockets of cells—often too small to be seen on magnetic resonance imaging (MRI)—can be found in margins around the tumor boundaries indicated by medical scans [5, 6]. The current standard for therapy involves either targeting the central, highly-visible tumor, a technique that allows the tumor to recur, or the expansion of surgical and radio-treatment area by a uniform margin [5, 7]. This “margin” approach is not specific to a patient’s brain geometry and results in over-treatment of healthy tissue, leading to cognitive decline and reduced quality of life [8]. Furthermore, these margins ignore the irregular growth of GBM tumors, which have varying growth rates depending on the tissue environment [9]. This leads to inadequate treatment in certain areas even when the margin is extended. Conversely, with spatiotemporal-aware tumor concentration approximation, treatment areas can be refined for recurrence-prone pockets and the preservation of healthy tissue. Thus, improving GBM tumor modeling is an area of particular interest towards effective, personalized neurosurgery and radiotherapy treatment planning.

The inhomogeneous growth patterns of GBM tumors are not only a consequence of cellular biology but also biomechanical dynamics such as solid stress caused by non-fluid tumor components [9]. These factors lead to mathematical tumor growth models being better estimates of cell infiltration than rudimentary human-defined margin approximations [5, 10]. The partial differential equation (PDE) is integral to many models [5, 6, 10–13] as it accounts for multiple variables: tumor cell infiltration and proliferation. The accepted paradigm of PDE-based brain tumor modeling is the Fisher-Kolmogorov Reaction-Diffusion Model [14]. This approach maps a tumor density PDE across brain geometry on an Eulerian framework:

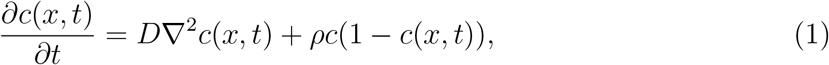

where *c, D*, and *ρ* represent tumor cell-density at location *x* and time *t*, tumor infiltration or diffusion coefficient, and cellular proliferation rate, respectively. The tumor’s growth is modeled forward to time *t* using numerical methods. However, the proposed PDE-based and other mathematical models have three main problems: 1) full analysis or even approximations of tumor growth can be very computationally expensive, requiring tens of thousands of simulations [5]; 2) the models do not consider whole brain geometry and only include a few human-defined features such as cell density and proliferation rate [11]; 3) many PDE-based models are restricted to an initial condition defined by a one-dimensional starting coordinate (tumor seed) rather than a volume of the patient’s current tumor. These issues have largely prevented the incorporation of tumor models in clinical settings.

Extensions of the Fisher-Kolmogorov model to computational, image-based solvers—such as Lipkova et al.’s state-of-the-art GliomaSolver [5]—that perform voxel-by-voxel calculations for cell density take several hours for just one-thousand forward simulations (predicting future tumor growth)^1^. Even with modern computing, execution times in the minutes for numerical solvers [11, 12, 15] are infeasible in practice because it requires a prohibitively long time (several days or weeks) to run tens of thousands of forward simulations. Due to the aggressive growth patterns of GBM [11], waiting weeks for computation allows significant changes in tumor features and volume, rendering the modeling futile. Treating a patient earlier would result in better outcomes for survival rate and quality of life [16]. Further, these simulations neglect the variability of anatomical boundary conditions between patients beyond simple tissue domain variability [17]. In Equation (1), *D* represents one of three different possible coefficients of tumor diffusivity:

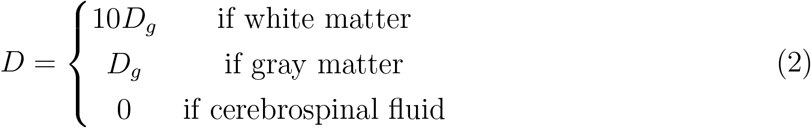

where *D*_*g*_ is the tumor diffusivity in gray matter [18]. Such models only incorporate a numerical change in tumor diffusion coefficient for various brain tissues, neglecting the wealth of additional anatomical information in MRI [11]. In other mathematical models [7, 19, 20], cell invasion in the brain is assumed to be both homogenous (uniform throughout all tissue) and isotropic (uniform in every direction); a tumor’s motility is represented as a scalar quantity rather than obtaining a deeper spatiotemporal understanding of tumor dynamics.

Deep learning has demonstrated great promise in the medical field for personalizing patient care [21, 22]. Neural networks have been successfully used for brain tumor segmentation and detection, validating the feasibility of extracting features from tumors and surrounding tissue [23, 24]. Notwithstanding, neural network literature has been sparse in the brain tumor-solving space, with few networks explicitly built for tumor modeling. A big-data approach to deep tumor modeling would require a significant amount of time-series brain imaging. In order to model unhindered tumor growth over time, longitudinal pre-treatment data is also required. However, such data is unavailable because most GBM patients are treated immediately after diagnosis [13]. Instead, creating a neural surrogate model^2^—using mathematical tumor growth simulations as ground-truth data—could offer solutions to the computational infeasibility of PDE-based tumor modeling while offering a proof-of-concept for bio-physical and patient anatomical learning.

Neural surrogate models—trained using mathematical evolutions as a ground-truth—in tumor modeling literature have been sparse and have only recently been proposed. Martens et al. utilize tumor concentrations at two time-steps as input data which is not clinically feasible due to treatment planning being conducted immediately after diagnosis [3, 25]. This model is also restricted to evolving tumors forward from a set time *t* = 0 as defined by the training data; its applicability to patients with varying initial tumor volumes is limited, and significant error compounds in the forward simulation by the time it evolves to the patient’s current tumor volume. More specifically, the model is not trained to start evolution at tumors of larger sizes, which is the input for most patients. In work by Ezhov et al., the network ignores the location of the patient’s tumor entirely and evolves the tumor from the center of the scan, starting from a one-dimensional point at time *t* = 0 [17]. This approach neglects learnable features from the patient’s tumor and is subject to the aforementioned clinical applicability and error compounding concerns. Thus, there is a need for approaches that can address the limitations of PDE-based solvers but can evolve tumors forward from numerous points in time and learn varying initial conditions (i.e., tumor volumes and locations).

In deep learning, the encoder-decoder with convolutional neural networks (CNN) has dramatically improved image-to-image translation—quickly becoming a standard architecture [26]. For image-to-image tasks, the encoder projects data to a lower dimension, which the decoder then upscales back to the original dimension. Its ability to “extract features” by bringing input data into the latent space (an abstract, internal encoding), coupled with a regression-based decoder, allows *n*-dimensional prediction. When applied to three-dimensional brain imaging, the convolutional operations within the encoder can learn spatial and geometric features, capturing both local and global contexts within an image [27]. The extracted spatial features from an input brain anatomy can inform a decoder that regenerates brain anatomy at a future state from the latent space. Further, these extracted features can allow the model to predict tumor growth in multiple directions and can be generalized to any spatial conditions and many points in time.

In this study, a deep encoder-decoder model is trained on glioblastoma patients and their corresponding mathematical simulations of future tumor growth. We propose a neural surrogate, GlioMod, that is two orders of magnitude faster than state-of-the-art PDE-based solvers. Our surrogate has applications for real-time treatment planning and is generalized to provide an understanding of each patient’s brain geometry. GlioMod serves as a method for tumor progression learning that can eliminate numerical solving altogether when timeseries, pre-treatment patient imaging becomes available. Furthermore, to the best of our knowledge, this network-based approach is the first in computational pathology to predict both brain tumor cell density at a future time step and also reconstruct the surrounding patient anatomy. The encoder built in this model can be utilized to extract valuable biophysical features to construct deeper characterizations of gliomas—for instance, to solve the tumor source localization problem [28]. The resultant model and simulation training data are provided as an open-source package at https://doi.org/10.5281/zenodo.6941367.

## 2 Methods

### 2.1 Data Curation and Masking

We use a dataset released by Lipkova et al. with eight glioblastoma patients [5]. This dataset includes T1-weighted (T1Gd) patient imaging—known to help reveal tumor cell density—along with corresponding white matter, gray matter, and cerebrospinal fluid segmentations [29]. Figure 1 depicts the various imaging modalities retrieved from the dataset.

**Figure 1:**
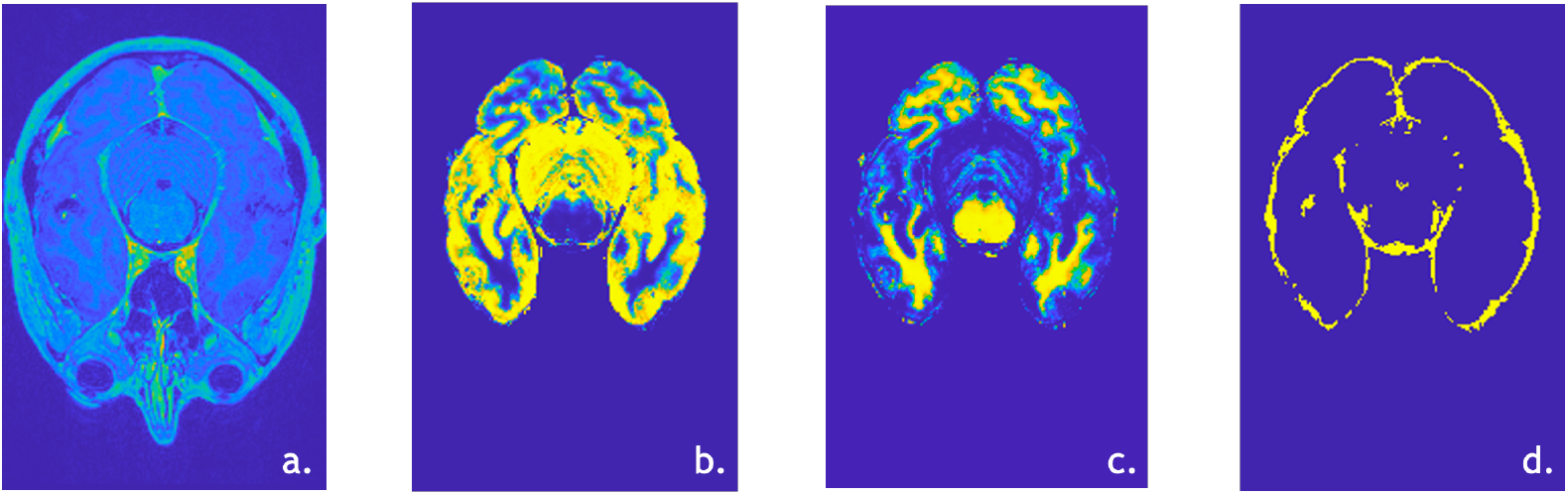
The imaging masks retrieved from 8 glioblastoma patients in order to form dataset. a) T1Gd scan b) Gray matter c) White matter d) Cerebrospinal fluid

The Neuroimaging Informatics Technology Initiative (NIfTI) formatted scans and masks are converted to a generic data file (DAT format); this is compatible with the numerical tumor solver used in Section 2.2 to generate the ground-truth tumor evolutions. The resultant DAT-formatted masks are padded with zeros to achieve 256 × 256 × 256 dimensions for uniformity and compatibility with the numerical solver.

#### 2.1.1 Synthetic Tumor Generation

In order to ensure a high number of training samples for deep learning, synthetic tumor locations were generated at randomized points in the patient anatomy. This serves to aid the neural model in learning tumor growth in a multitude of anatomical conditions. The numerical solver in Section 2.2 generates future tumor density simulations from these one-dimensional points. The data for two of the eight patients were discarded due to overestimated cerebrospinal fluid volumes, which resulted in synthetic tumors with no growth due to zero tumor diffusivity within this fluid. One hundred synthetic tumor locations were generated within the brain geometries of the six patients whose data was kept. The randomized points, “tumor seeds,” are generated within a NIfTI mask that combines the white and gray matter volumes. Seeds sufficiently close to the brain boundary are discarded due to zero diffusivity outside the brain. In order to make this restriction, an approximation is made of the central brain volume based on averages of the patient data. This binary mask of white and gray matter is depicted in Figure 2.

**Figure 2:**
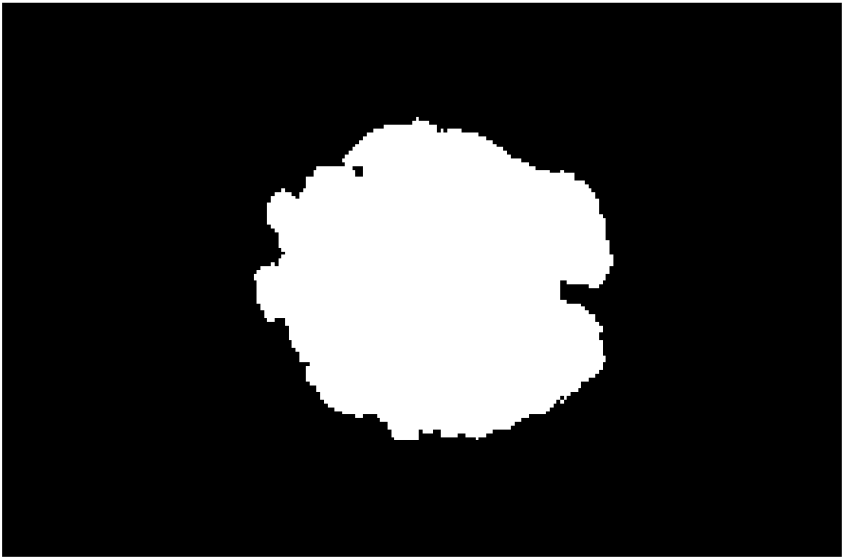
The central brain volume approximation utilized to restrict boundaries in which synthetic tumor seeds can be generated.

Additionally, tumor seeds are not generated within cerebrospinal fluid due to its zero diffusivity, resulting in no tumor growth. Examples of tumor seed generations with these restrictions are depicted in Figure 3.

**Figure 3:**
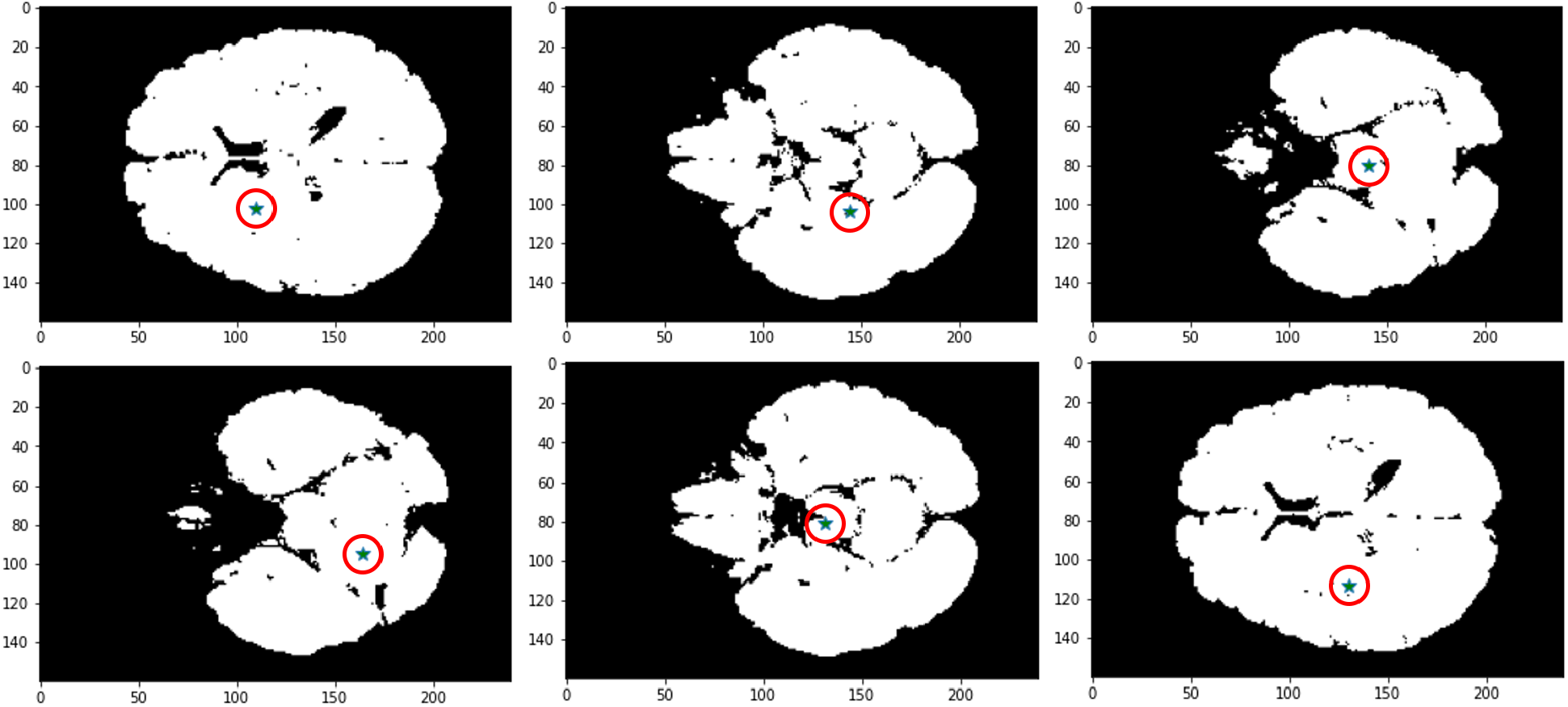
Montage of tumor seeds generated in XY-plane at varying Z values. Teal asterisk (*\**) represents the tumor seed location (*x, y, z*) where *x ∈* [0, 240], *y ∈* [0, 160], *z ∈* [130, 170].

Tumor seed coordinates are normalized to [0, 1] with respect to the image size to serve as input to the mathematical solver.

### 2.2 Numerical Tumor Solving with Reaction-Diffusion

Lipkova et al.’s GliomaSolver is utilized to evolve brain tumors forward in the six patient anatomies at the tumor seeds generated in Section 2.1.1. It accepts input segmentation masks of gray matter, white matter, and cerebrospinal fluid. GliomaSolver outputs a simulated tumor cell concentration until day *t* at ∆*d* day intervals. The parameters utilized for tumor simulation are specified in Table 1.

**Table 1:** Input specifications to numerical solver for tumor evolution. (1) In the left-hand column, static inputs to the GliomaSolver tool are listed. (2) In the right-hand column, their respective settings are given.

| Tumor Parameters | Value |
| --- | --- |
| White matter diffusivity ($D_w$) | $0.0013 \left[ \frac{cm^2}{day} \right]$ |
| Gray matter diffusivity ($D_g$) | $0.1 \times D_w$ |
| Cellular proliferation coefficient ($\rho$) | $0.025/day$ |
| Days to evolve forward ($t$) | 500 |
| Day interval to dump ($\Delta d$) | 50 |

GliomaSolver’s reaction-diffusion tumor growth model is utilized to generate 100 synthetic tumors at the locations of the tumor seeds from Section 2.1.1. A total of 6,000 data points are retrieved from simulations via GliomaSolver, taking seven hours on a machine with specifications listed in Appendix A.1.

The solver gives Visualization Toolkit Unstructured Grid (VTU) formatted files as output. This base64-encoded unstructured grid is specific for visualization in the ParaView software. The VTU grids have five different image channels, each representing different tissues. Channel one is of particular interest because it includes both tumor cell concentration and surrounding brain anatomy. Figure 4 depicts such outputs in ParaView as predicted by the mathematical solver.

**Figure 4:**
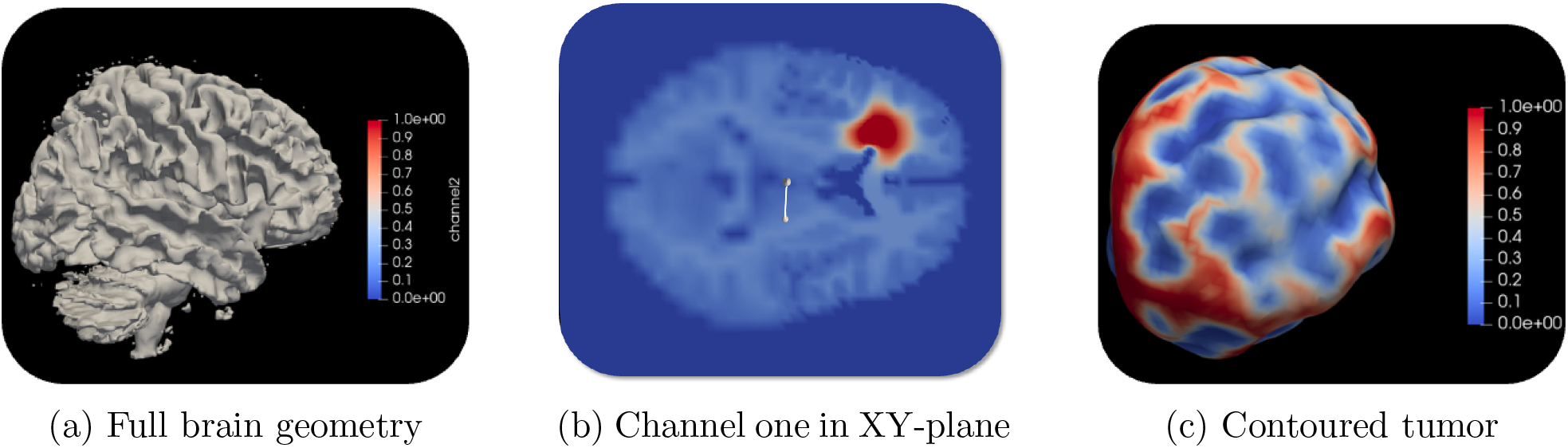
Visualizations of brain geometry and tumor concentration outputs from mathematical solver. Tumor is simulated to time *t* = 500 days.

Although this unstructured grid is functional for visualizations, it is not ideal for deep neural modeling via convolution^3^. An in-house Python script is utilized to cast the VTU mesh as tabular data. The tabular data consists of four columns with the (*x, y, z*) coordinate and this point’s value of channel one, a quantity represented by Equation (3):

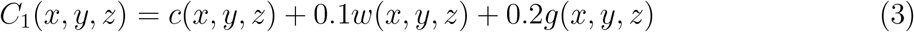

where, for every point, *C*_1_ represents the value of channel one, *c* represents tumor presence, *w* represents white matter presence, and *g* represents gray matter presence. The in-house script also deletes points at which *C*_1_ is zero to reduce data size. Figure 5 depicts the tabular data with and without the redundant zeros.

**Figure 5:**
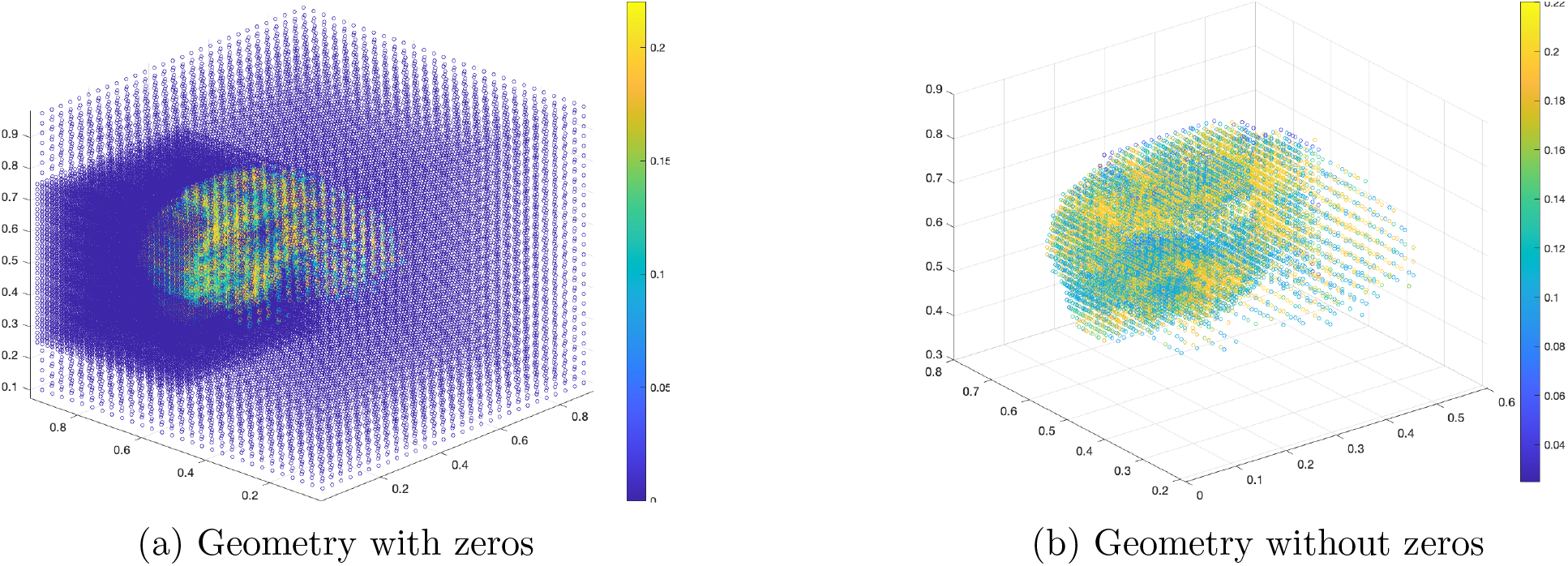
Scatterplots depicting brain geometry of channel one output from mathematical solver in a three-dimensional space.

In intermediate forward evolution dumps, the mathematical solver does not fully render the brain geometry. Rather, it keeps the local tumor area in high resolution and jumps resolution in the surrounding area. This inadvertent feature of the training data is deemed helpful for neural solving in Section 2.3 because it sets an attention area over the voxels nearest to the geometry that directly affects tumor diffusivity and infiltration into surrounding tissue. This attention area is most visible in Figure 6 which depicts a comparison of the initial brain geometry and the tumor evolution at a later time-step.

**Figure 6:**
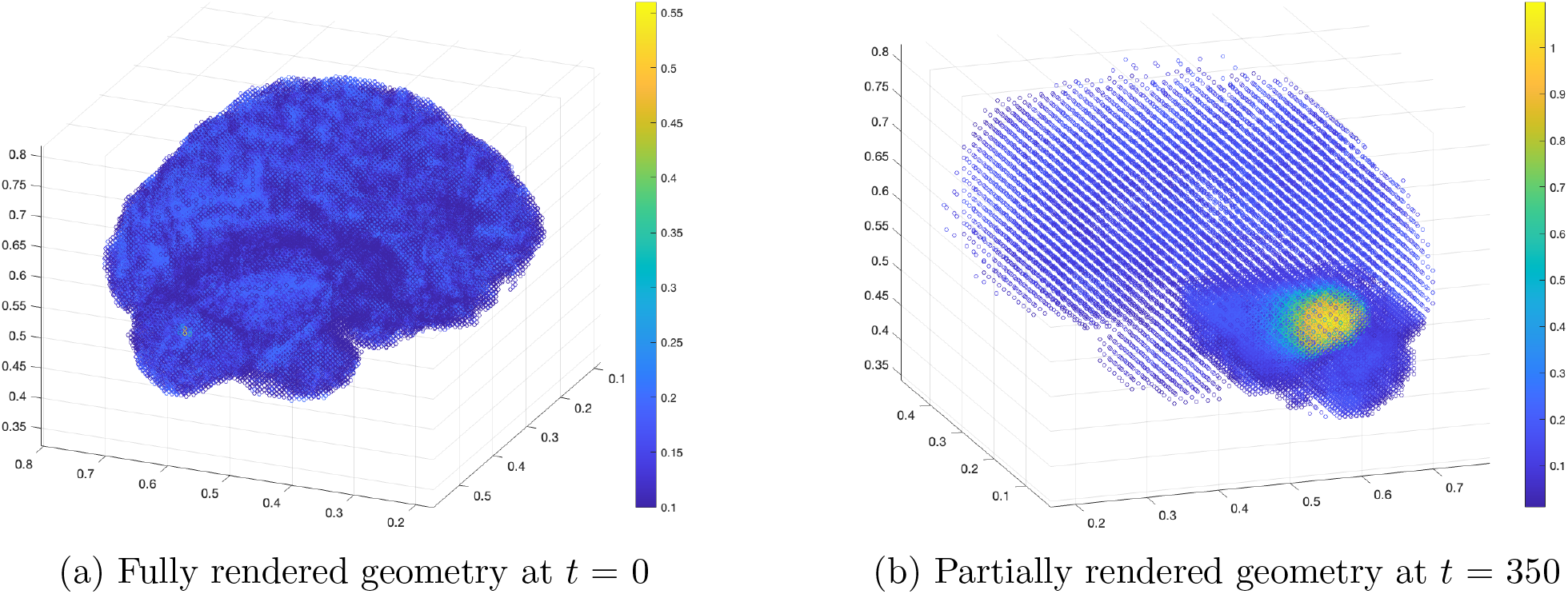
Mathematical solver renders patient geometry far away from the tumor in a lower resolution in order to accelerate computation and prioritize regions of interest.

### 2.3 Deep Learning for Tumor Evolution

#### 2.3.1 Data Interpolation & Preparation

For a convolution operation to extract valuable features, data must be arranged such that the features are spatially ordered in the dataset. In GlioMod, there are time-series images with predictions ordered in time. However, exploiting the spatial properties of the images requires a different setup than the tabular data from Section 2.2. Hypothetically, a 1 × 3 convolution kernel can be applied to tabular data, but this would first consider data (*x, y, z*) then (*y, z, c*), which is not conclusive for spatial understanding. The tabular data must be transformed into a four-dimensional matrix to which convolution filters can be applied. The four dimensions are (i) height, (ii) width, (iii) depth, and (iv) channels. Interpolation to a 256 × 256 × 256 matrix is ideal; however, in practice, the compute required to load the matrix exceeds capacity (requires over 40 gigabytes of RAM). Therefore, we interpolate a 64 × 64 × 64 matrix by normalizing each (*x, y, z*) point from 0 to 64 and setting that point equal to the value of channel one. The resulting matrix has a single channel because it is grayscale. An example of patient geometry at this new size is depicted in Figure 7.

**Figure 7:**
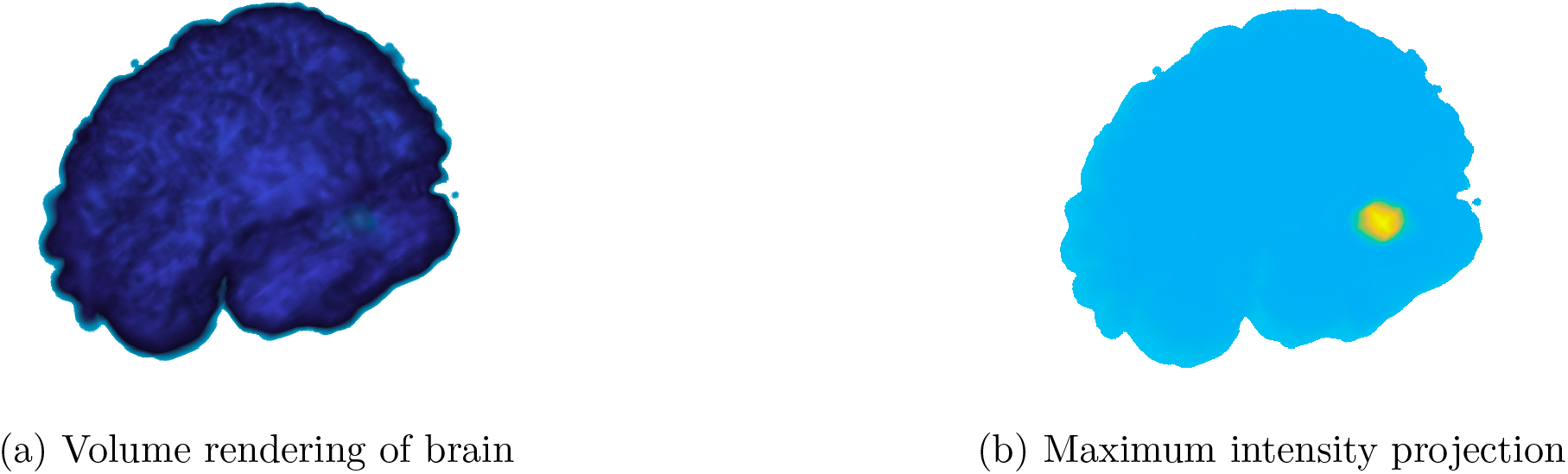
Visualization of brain geometry interpolated to 64 × 64 × 64 size at time *t* = 450 accompanied with a maximum intensity projection highlighting tumor concentration.

Of the six patients and 6,000 corresponding total synthetic tumors, one patient is withheld for testing the generalizability of the model on unseen brain geometries. With 5 patients × 100 tumors per patient × 10 initial starting times, there were 5,000 remaining samples that encompassed days 0 through 500. From these samples, two four-dimensional matrices each with 4,500 samples are constructed: 1) input matrix with input samples ranging from day 0 to 450, and 2) output matrix with response samples ranging from day 50 to 500. Because the encoder-decoder model has a fixed depth, the training data must have a set interval that the model learns to evolve forward. The constructed matrices facilitate model training such that the model is always predicting channel one (Equation (3)) 50 days forward, after the current input. For instance, a training input could be the channel one image at 250 days, and its ground-truth output is the evolved image at 300 days—as solved in Section 2.2.

#### 2.3.2 Model Architecture & Hyperparameter Optimization

Predicting future tumor states requires an understanding of bio-physical properties and patient brain geometry. In order to better assimilate these properties and capture underlying spatiotemporal features compared to PDE-based approaches, deep learning is utilized. GlioMod uses a novel encoder-decoder neural network which performs image-to-image regression to predict future tumor states. This architecture is depicted in Figure 8.

**Figure 8:**
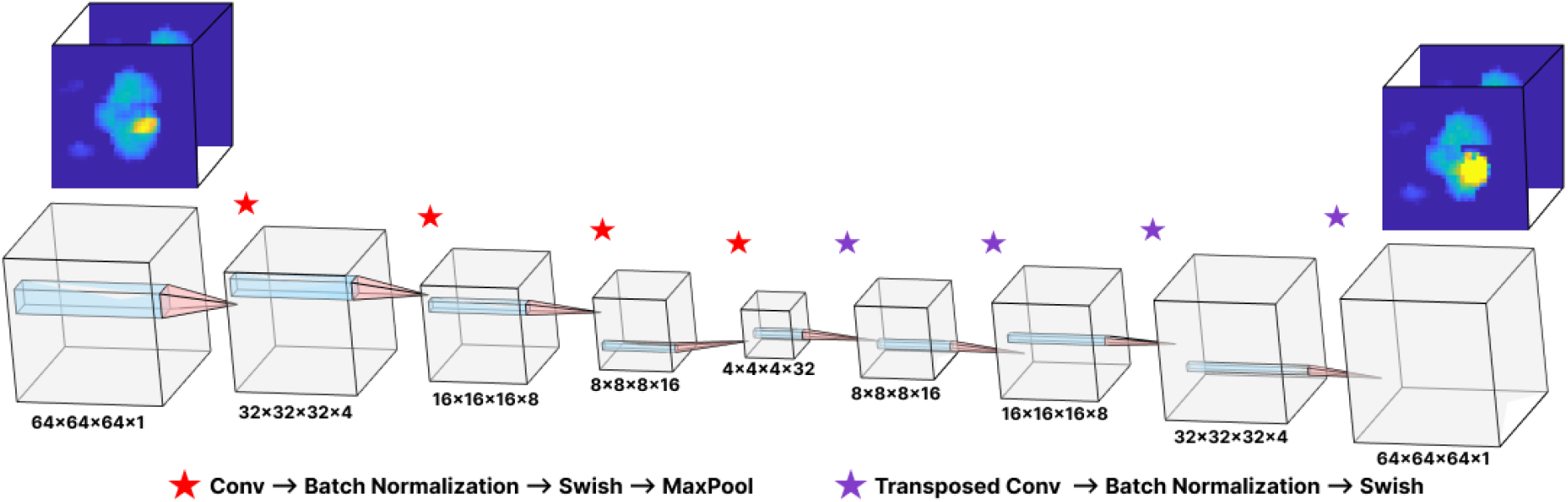
Diagram of 33-layer deep encoder-decoder network architecture with 36,500 learnables. 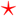 represents 3D Convolution ⇀ Batch Normalization ⇀ Swish ⇀ 3D Max Pool operations. 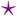 represents Transposed Convolution ⇀ Batch Normalization ⇀ Swish operations.

The architecture was iteratively designed based on the training results after initially adopting a 19-layer deep network.

Primary changes in architecture include the utilization of Swish activation as opposed to Rectified Linear Unit (ReLU), implementation of batch normalization, and the adoption of larger kernel sizes in initial convolutions. The Swish function has been observed to consistently outperform ReLU but is more computationally intensive [30]. Batch normalization was implemented into the encoder-decoder design between the convolution and nonlinearity to help the model converge faster and reduce training time. Larger kernel sizes such as the 7 × 7 × 7 filter in the first convolution followed by the smaller 3 × 3 × 3 filters later in the model take both greater tumor surrounding context and tumor edge context into account.

The model hyperparameters were tuned iteratively based on model results on the training dataset. The iterative hyperparameter optimization methods prevent the possibility of fitting to data while maintaining high performance. The optimized hyperparameters used for model training are indicated in Table 2.

**Table 2:** Hyperparameter optimization is conducted to finetune model performance. In the left-hand column, 8 hyperparameters are listed. In the right-hand column, their respective settings are given based on the testing conducted in this study.

| **Model Hyperparameter** | **Value** |
| --- | --- |
| Epochs | 30 |
| Minibatch Size | 1 |
| Initial Learning Rate | 0.006 |
| Learn Rate Decay | 0.1 |
| Learn Rate Drop Period | 1 |
| L2 Regularization | 0.0001 |
| Momentum | 0.9 |
| Gradient Decay Factor | 0.9 |

The learning rate hyperparameter followed a piecewise schedule. It had an initial condition as indicated by Table 2, and was multiplied by the decay factor every epoch. This initial learning rate was deemed to have the best tradeoff between convergence and precise training. The decay was implemented to achieve better network generalization and improve the learning of complex patterns later in training.

## 3 Results

To validate the performance of GlioMod, we use multiple established error-based metrics for regression tasks such as root mean squared error, a quantitative and qualitative analysis of tumor prediction agreement on brain geometry, and model run-time. The final GlioMod neural model was trained for 30 epochs with a loss function of half mean square error. Its final training graph with this loss function calculation is shown in Appendix B.

### 3.1 Regression Error Metrics

For image-to-image regression tasks, models are commonly evaluated based on their error: a prediction’s deviation from the ground-truth intensity for each pixel in an image. Four regression error metrics are utilized to evaluate GlioMod: root mean squared error (RMSE), relative root mean squared error (RRMSE), and relative squared error (RSE). These metrics are calculated using formulae described in Appendix C. GlioMod simulates tumor growth on the test subset of 900 image pairs—with input and output ranging from days 0 to 450 and days 50 to 500, respectively. The predicted images by GlioMod are compared to the groundtruth, mathematically-predicted output samples. On the test subset (n=900), we find that GlioMod yields a 0.0204 *±* 0.0001 RMSE, 0.0013 *±* 0.00001 RRMSE, and 0.3735 *±* 0.0049 RSE. These error metrics are depicted in Figure 9.

**Figure 9:**
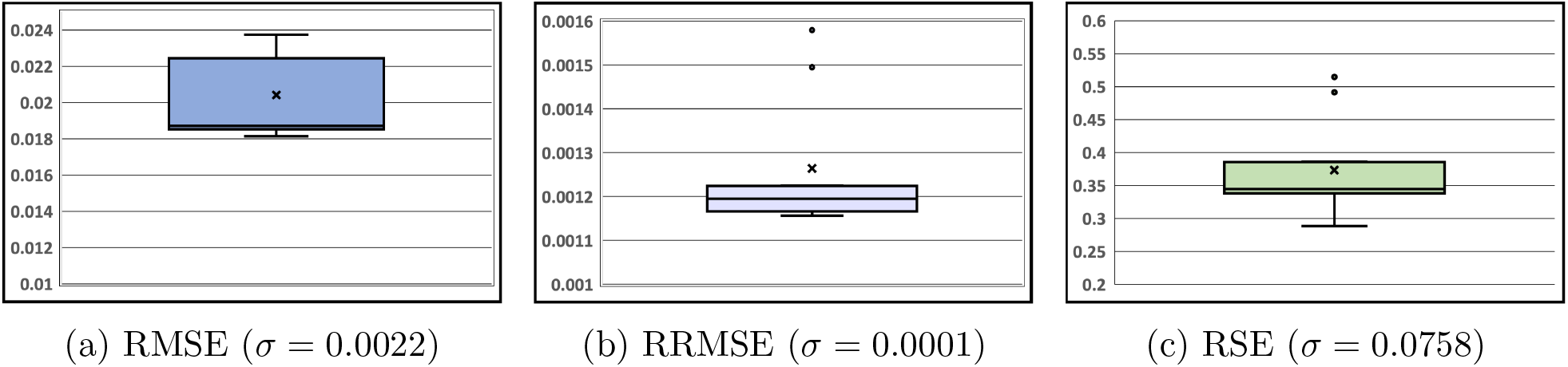
GlioMod shows low error on standard regression metrics (n=900).

Further, error is quantified by calculating agreement between GlioMod-predicted and ground-truth simulations on the testing dataset. Each *predicted* and *truth* simulation is condensed into a single value by summing the values of every pixel in the image. The predicted and truth values are paired, and linear regression is conducted on the 900 resultant paired points. To reveal systematic error in the model, Bland-Altman analysis is conducted, measuring agreement between GlioMod and ground-truth predictions. The agreement analysis, depicted in Figure 10, yielded an *R*^2^ value of 0.95 and raw error of *−*70.0.

**Figure 10:**
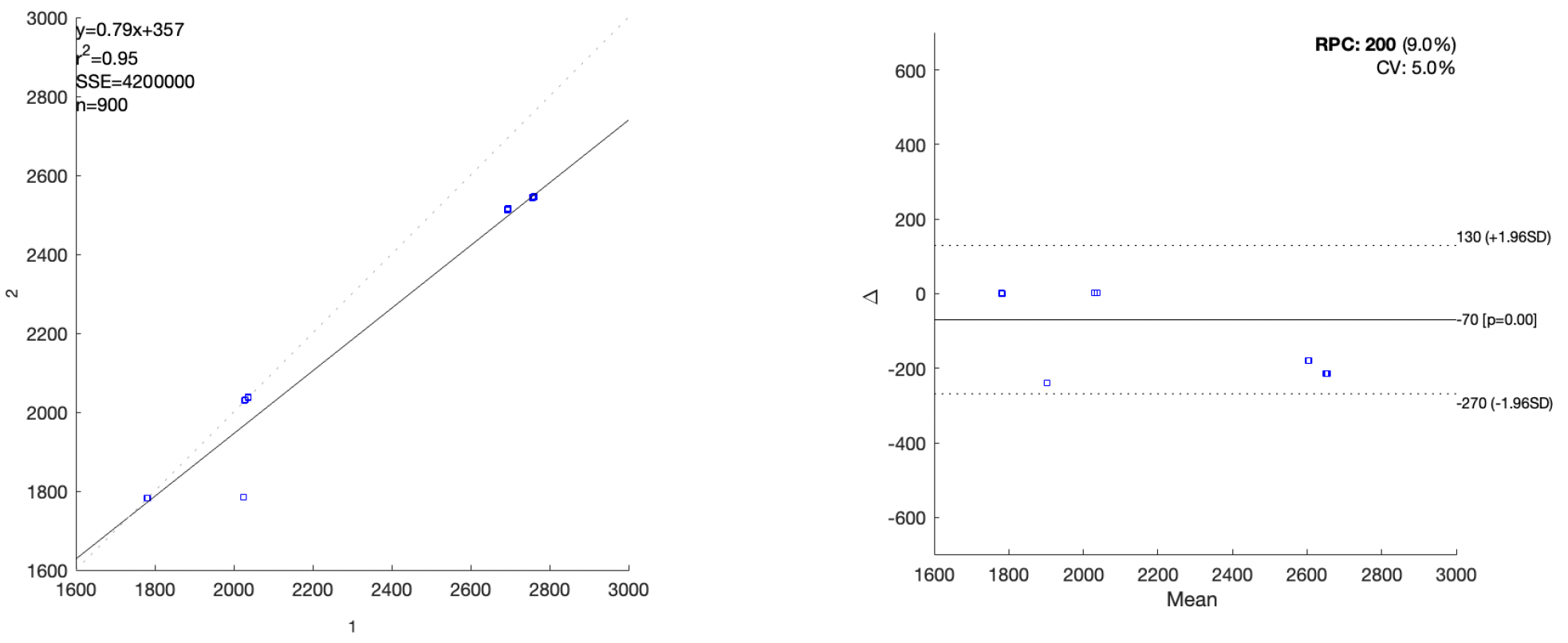
Analysis suggests high agreement between GlioMod simulations and corresponding ground-truth (n=900). (Left) Predicted simulation (Y-axis) is plotted versus ground-truth simulation values (X-axis) to yield line of best fit of *y* = 0.79*x* + 357. (Right) Bland-Altman analysis suggests systematic underestimation of channel one concentration.

### 3.2 Qualitative Analysis of Agreement

To visualize the simulation error in brain geometries, qualitative analysis of error on twodimensional slices of the brain anatomy is conducted. A montage of tumor values at varying time steps is displayed in Figure 11 which highlights the differences in the neural solver to ground-truth evolutions.

**Figure 11:**
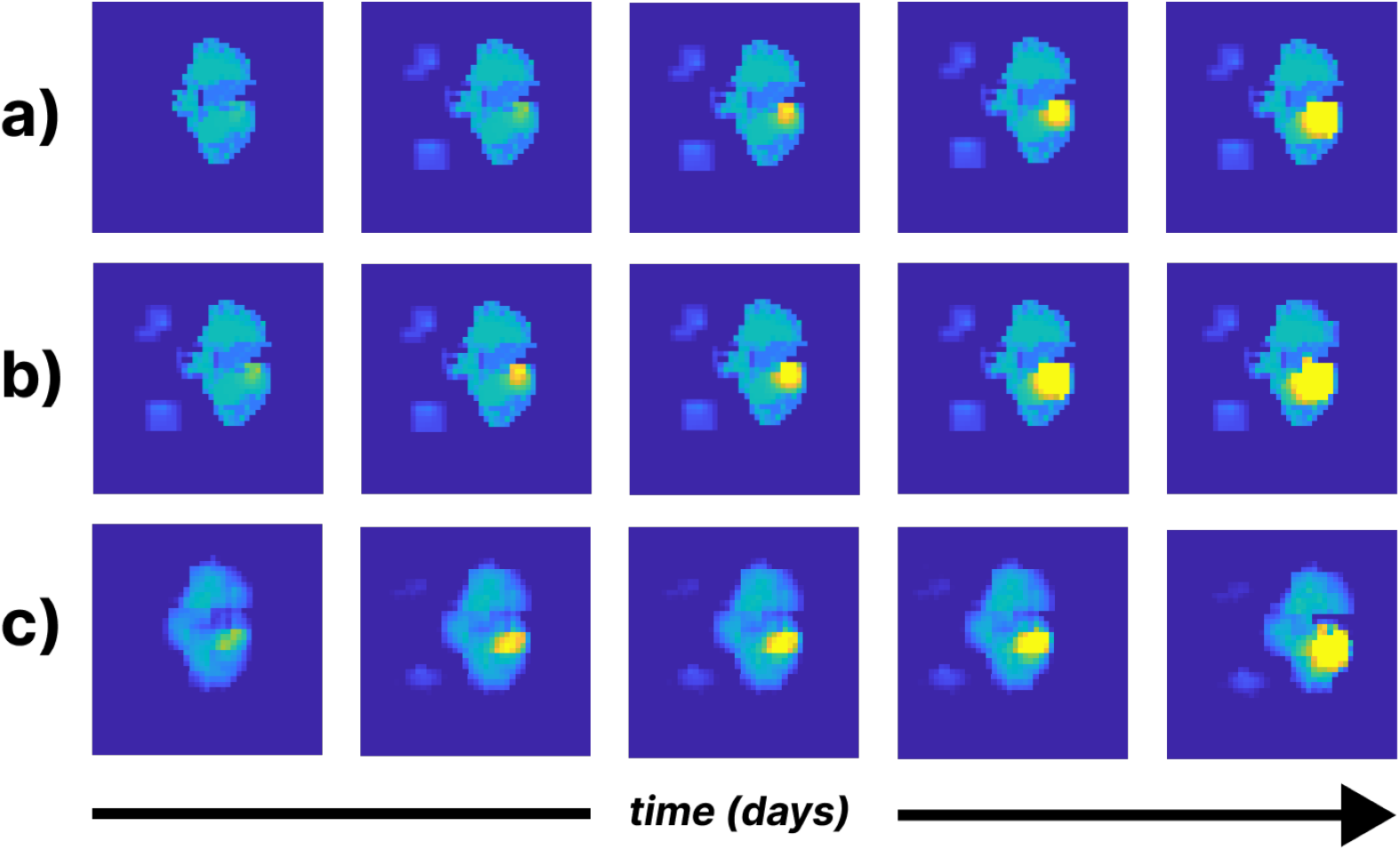
Montage of XY-plane slices of tumor concentrations and brain volumes at 50 day intervals. a) Input from day 250 to day 450; b) Ground-truth, GliomaSolver predictions from day 300 to day 500; c) Neural model predictions from day 300 to day 500.

While predicting tumor concentration is the primary endpoint of this study, reconstructing surrounding patient geometry is an important measure of model success. This is quantified in the error analysis of Section 3.1 and qualitatively shown in Figure 12 which depicts the reconstruction of patient geometry in a forward timestep despite having tumor growth.

**Figure 12:**
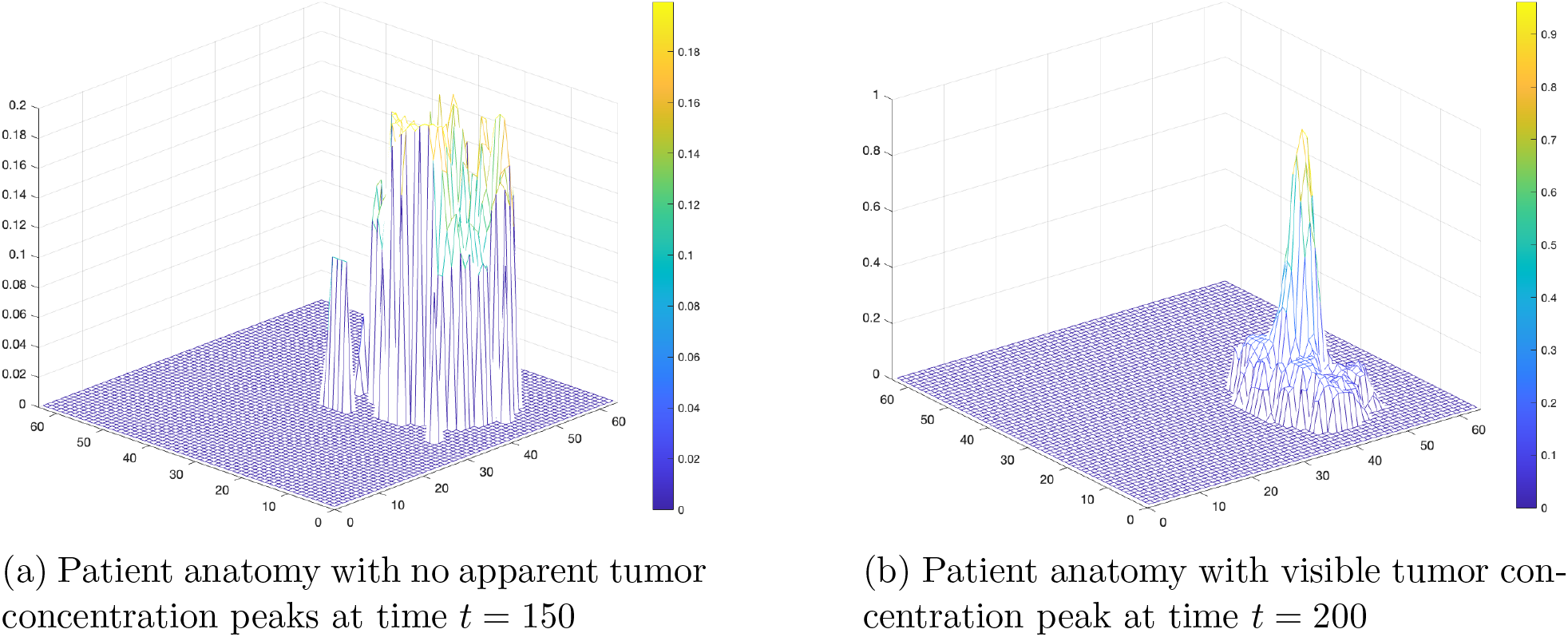
GlioMod reconstructs surrounding patient geometry while growing tumor.

**Figure 13:**
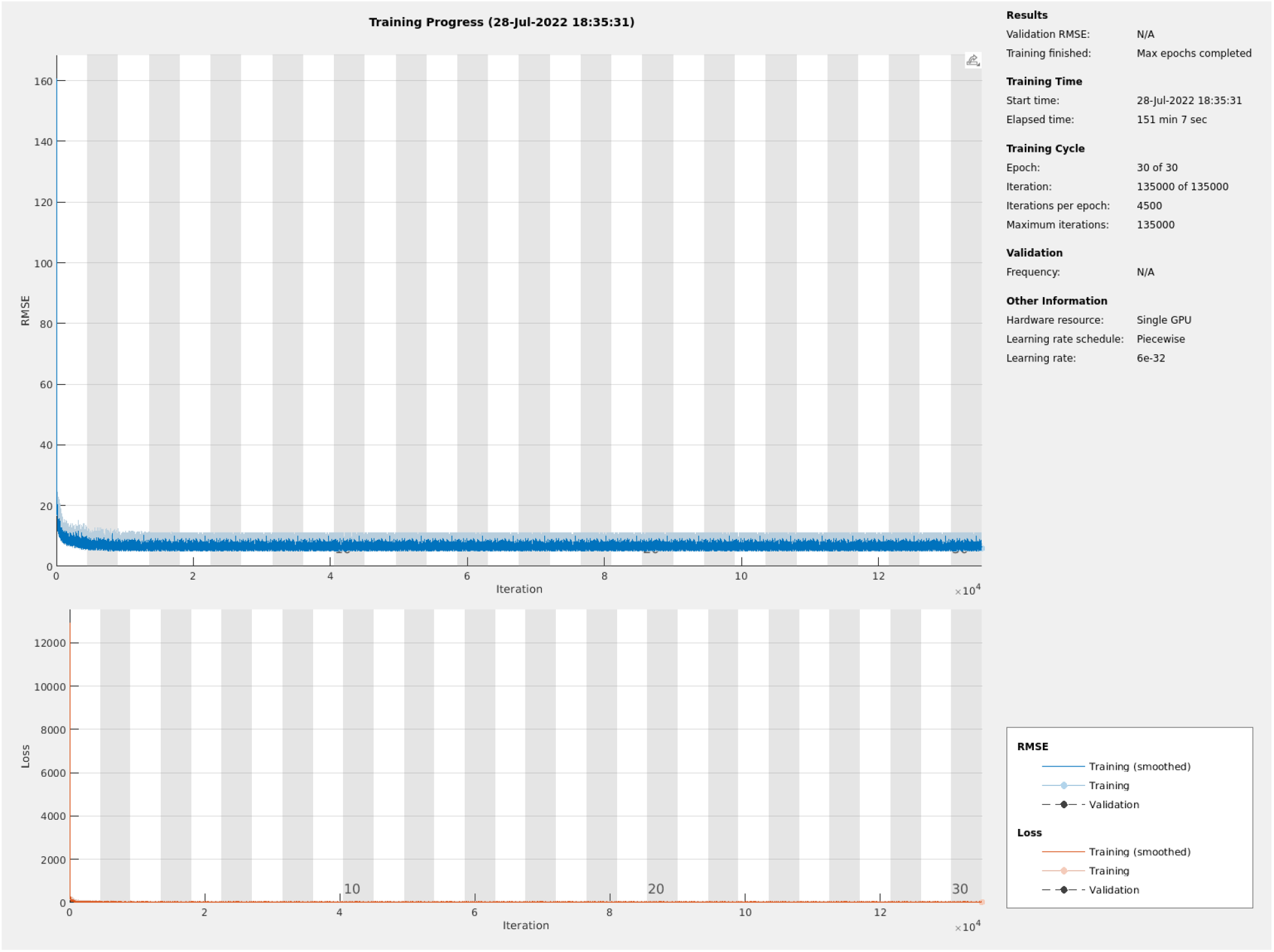
Progress over 30 epochs of training GlioMod. Top graph depicts training root mean square error plateau and bottom graph depicts half mean square error plateau.

### 3.3 Prediction Run-time

Using a testing system as described in Appendix A.2, simulation times for different tumor growth modeling methods were measured on the testing dataset. run-times are based off of each patient requiring 1,000 tumor simulations, as is currently needed for probabilistic modeling, and each simulation requiring 10 simulation outputs until time *t* = 500 days. The time per patient is extrapolated from the actually measured time per simulation. The scores are shown in Table 3.

**Table 3:** GlioMod run-time is clinically relevant with significantly faster run times compared to state-of-the-art approaches. Measurements for GlioMod and GliomaSolver are averages of three trials while PDE-based frameworks and Lattice Boltzmann run-times come from past literature [11, 12, 15, 17].

| Approach | Time Per Patient | Time Per Simulation |
| --- | --- | --- |
| GlioMod | $\sim 0.47$ hours | 1.684 seconds |
| GliomaSolver | $\sim 30.69$ hours | 110.5 seconds |
| PDE Frameworks | $\sim 55 - 111$ hours | $\sim 200 - 400$ seconds |
| Lattice Boltzmann | $\sim 200 - 250$ hours | $\sim 750 - 900$ seconds |

## 4 Discussion

In this paper, we find that spatiotemporal-aware brain tumor modeling achieved via deep learning is an effective technique for tumor concentration prediction and anatomical reconstruction. While previous research in neural modeling of GBM tumor growth is not personalized and neglects a patient’s current tumor volume, our model is specific to each patient’s geometry. Using this approach, we find that our resultant model, GlioMod, is capable of reproducing a realistic tumor growth pattern and concentration in close agreement (*R*^2^ = 0.95) to state-of-the-art mathematical solvers.

In Sections 2.1 and 2.2, a unique synthetic tumor generation method is described in which tumors are grown in a variety of input geometries. This step was necessary due to the unavailability of longitudinal pre-treatment data for GBM patients. Although the synthetic tumor generation method used in this work does not perfectly match real tumor growth behavior—as existing mathematical models are imperfect—it serves as a reasonable surrogate and proof of concept. A limitation of the proposed generation method in Section 2.1.1 is the tendency for 95% of GBM tumors to manifest in the supratentorial region of the brain [31]. Although the majority of tumor seeds were indeed in this region, this distribution could be explicitly controlled, improving the applicability of GlioMod to real patient tumors. The ability to train the model on a wide range of brain geometries and patient conditions is critical to the success of any prediction model. The partial-rendering data input described in Section 2.2 encourages the model to focus on the local tumor region. This takes full advantage of the data, given that the ground-truth is based on a mathematical model that only considers the variation of nearby tissues. However, it is noted that given longitudinal data of real tumor growth, the partial-rendering technique should not be used to encourage the model to make predictions based on full brain renderings.

In order to train the deep neural network, one of the six patients is withheld to be used as testing data. This approach was preferred as opposed to withholding 10% of the samples from every patient. In our experiments, we note that the model had a slight bias towards the training geometries, even if it had not seen specific testing seeds. Thus, using an utterly unseen brain geometry allows us to properly gauge the performance of the model. This research proposes a novel encoder-decoder architecture that can be utilized for GBM imaging. In the final architecture, we utilize the Swish activation and Batch Normalization functions. However, I also tested ReLU activation with and without normalization. Swish offered approximately a 2% improvement in training loss over ReLU. We also observe the importance of batch normalization in allowing the model to converge independent of learning rate. Experimentally, batch normalization enabled the use of a higher learning rate which allowed the model to converge and achieve better performance. Without this implementation, the model would underfit at a loss plateau and consistently predict a general brain geometry.

In the statistical data analysis, we use RMSE, RRMSE, RSE, linear regression, and Bland-Altman analysis to quantify the performance of GlioMod. The regression error statistics indicate low error and excellent performance for predicting tumor concentration and reconstructing anatomy. Although our analysis controls for the importance of the primary task—tumor cell concentration prediction—by weighting it over brain geometry (see Equation (3)), no quantitative analysis is conducted solely on tumor concentration. Thus, a deeper analysis limited to a subset of pixels with intensity over 0.3 (to focus on tumor concentration) may indicate specific characteristics of model prediction on tumor concentration.

Linear regression revealed a strong *R*^2^ value of 0.95, suggesting a close fit and agreement between the mathematical model and neural surrogate. In the Bland-Altman analysis, a systematic mean error of *−*70 units is identified. This is within approximately 3.45% of the total value in the images, indicating excellent performance. One characteristic of interest in both the linear regression and Bland-Altman analysis is the clustering of points around the plane. Although there are 900 points, there appear to be only six distinct clusters of predictions. This is likely because the model makes the same type of errors independent of location depending on the timestep of tumor evolution. It is observed that the error is larger for tumors of higher ages. Because of the close agreement to the mathematical model yet distinct qualitative variations in tumor progression, it is likely that GlioMod is more generalized to patient anatomy based on the features present in imaging.

The trained GlioMod neural network model can be accessed through an open-source software package at https://doi.org/10.5281/zenodo.6941367. Finally, to increase clinical feasibility, GlioMod can be incorporated into treatment planning software to give real-time suggested treatment margins for a patient.

## 5 Future Work

The *black-box* characteristic of deep learning results in interpretability challenges compared to the many proposed bio-physics models. Neural ordinary differential equations (ODE) are a new family of neural network models [32]. These models not only have greater interpretability compared to traditional neural network-based approaches but also an ability to contextualize localized behavior within a dataset [33]. A neural ODE can be implemented to evolve latent space features before the decoder module. With a continuous-depth model, evolution can be integrated over any period of time (i.e., 314.5 days) allowing arbitrary combinations of input data.

Currently, the model is trained on 64×64×64 images due to high resource requirements of higher resolution 3D images. One approach to reducing data size while maintaining a higher resolution is to identify a region of interest on each brain scan and train on this region only. The mathematical solver utilized to generate the ground-truth tumor concentrations in this study has its own method to render far-away voxels in a lower resolution to reduce computational cost. A similar strategy can be extended to model training. This adaptive resolution approach suffers from the drawback of losing some global context in the image.

Because nearly all glioblastoma patients receive treatment, pretreatment time-series scan data is unavailable. However, if the tumor modeling problem is expanded to include chemotherapy and radiotherapy, the model will have access to additional information regarding the tumor response over time and the effects of treatment. The model may be trained to predict tumor growth over the course of a whole treatment plan. Future work into a multi-parameter model that considers tumor shrinkage due to chemoradiotherapy will provide oncologists with an even more clinically-applicable tool.

## 6 Conclusion

In this study, a glioblastoma multiforme tumor evolution model, GlioMod, is proposed to predict future glioma tumor growth using an encoder-decoder convolutional neural network. This surrogate model has the ability to learn spatiotemporal features of tumor growth and brain geometry—which help inform better treatment planning. we generated 5,000 synthetic tumors in real patient brain anatomies using PDE-based modeling, resulting in 4,500 paired-MRI states for training. In order to validate GlioMod, 900 pairs of input patient anatomies and their corresponding PDE-solved future tumor concentrations are evaluated against GlioMod’s predictions. GlioMod has an RRMSE of 0.0013 *±* 0.00001, suggesting excellent performance on the task of reconstructing tumor anatomy and tumor concentration. The results indicate that GlioMod can predict tumor growth with a high accuracy and close agreement to mathematical modeling while being 2 orders of magnitude faster. GlioMod serves as a validated method for tumor growth learning that can replace mathematical solving when longitudinal patient imaging becomes available. Furthermore, it enables the use of tumor models in the clinic by being specific to every patient’s unique brain geometry and glioma conditions. In the next stage of research, GlioMod can be extended to a neural ordinary differential equation framework in which continuous predictions can be made.

## Data Availability

All data produced in the present study are available upon reasonable request to the authors.

https://doi.org/10.5281/zenodo.6941367

## A Machine Specifications

### A.1 Testing Machine

The benchmarking for optimization run-time was conducted on the testing machine with specifications below:

- CPU: 8-core Apple M1
- RAM: 8 Gigabytes RAM
- GPU: 8-core Apple M1

### A.2 Training Machine

The GlioMod model was built and trained on the MSEAS computing cluster with specifications below:

- CPU: 16-core Intel Xeon Gold 6142 @ 2.60GHz
- RAM: 73 Gigabytes RAM
- GPU: NVIDIA PNY RTX A6000

The software specifications on which the GlioMod tool was built are specified below, along with the environment setup for model training:

- CentOS Linux 7.9.2009
- MATLAB 9.11 R2022a
- Python 3.9 (64-bit)

## B Training Progress

## C Error Formulae

The following describe the regression error formulae utilized in Section 3.1 for validation and testing of GlioMod. Root mean squared error is calculated using Equation 4.

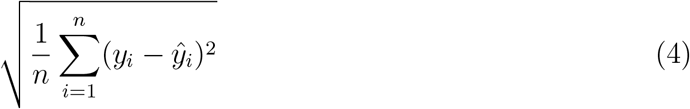

where *n* represents the number of predictions (i.e. pixels in an image), *y*_*i*_ represents the ground-truth value for the prediction, and *ŷ*_*i*_ represents the model predicted value.

Relative root mean squared error is calculated using Equation 5.

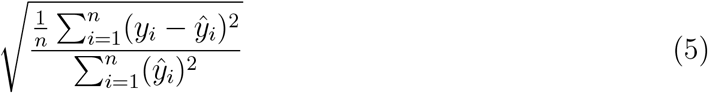

Relative squared error is calculated using Equation 6.

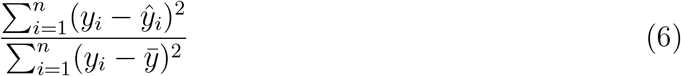

where 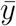 is calculated using Equation 7.

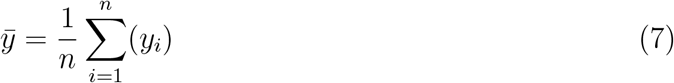

## Footnotes

1 On a machine with specifications as described in Appendix A.1.

2 A surrogate model in machine learning is where ground-truth data may not be available, so statistical or mathematical models are utilized to approximate a simulation output.

3 A technical explanation for this can be found in Section 2.3.

